# Association Between Device-Detected Obstructive Sleep Apnea and Atrial Arrhythmia Burden in Patients Monitored by HeartLogic-Enabled Cardiac Devices: A Retrospective Cohort Study

**DOI:** 10.1101/2025.09.09.25335413

**Authors:** Dylan Rajaratnam, Justin Tan, Kathryn Wales, Andrew Hopkins, Joseph Assad, Hany Dimitri

## Abstract

**Background:** Sleep-disordered breathing, particularly obstructive sleep apnea (OSA), has been associated with increased atrial arrhythmia burden in various populations (1). The Boston Scientific HeartLogic platform offers continuous, multiparametric monitoring and atrial arrhythmia burden. We aimed to evaluate the association between device-detected OSA severity and atrial arrhythmia duration in patients with implantable cardioverter-defibrillators (ICD) or cardiac resynchronization therapy defibrillators (CRT-D) enabled with HeartLogic.

**Methods:** We performed a retrospective cohort study of adult patients monitored via the Boston Scientific HeartLogic platform between January 2018 and December 2023. Patients without complete remote monitoring data or inactive HeartLogic indices were excluded. Obstructive sleep apnea severity was estimated using the AP Scan-derived Apnoea-Hypopnoea Index (AHI) equivalent. Atrial arrhythmia burden was calculated as the average daily duration of atrial high-rate episodes.

**Results:** Among patients who had atrial arrythmias, a significant positive association was found between increasing AHI and atrial arrhythmia duration. For every 1-unit increase in AHI, arrhythmia burden increased by approximately 10.55 seconds/day (p = 0.005). Compared to patients with no sleep apnea, those with Mild, Moderate, and Severe AHI had significantly higher daily arrhythmia durations compared to baseline by 2.8, 2.8, and 3.0 hours respectively (all p < 0.001).

**Conclusion:** Device-detected obstructive sleep apnea (OSA) was found to be associated with increased atrial arrhythmia burden in patients with cardiac implantable electronic devices (CIEDs) monitored by the HeartLogic platform. These findings generate important hypotheses regarding the integration of remote OSA surveillance into arrhythmia risk stratification and heart failure management and warrant further prospective investigation.

## Introduction

Obstructive sleep apnea (OSA) is a highly prevalent sleep-related breathing disorder characterized by repetitive episodes of upper airway obstruction during sleep, resulting in intermittent hypoxia and hypercapnia (1). OSA has been increasingly recognized as a key contributor to cardiovascular morbidity and mortality (2). OSA affects 21–82% of patients with heart failure depending on the severity of heart failure and up to 49% of individuals with atrial fibrillation (AF), establishing a strong and clinically meaningful bidirectional relationship (3).

In patients with heart failure, OSA leads to an increase in sympathetic activity, negative intrathoracic pressure swings, adverse haemodynamic changes, sleep fragmentation and fluid retention, all of which can exacerbate both left and right ventricular dysfunction (1). The cyclical nature of hypoxia and intrathoracic pressure swings also promotes structural and electrical remodelling of the atria, thereby contributing to the development and maintenance of AF. Several observational studies and mechanistic investigations have suggested that treating OSA, particularly with continuous positive airway pressure (CPAP) may reduce the recurrence of AF by 42%, and patients with untreated OSA have a 57% increased risk of AF recurrence (4). Sleep-disordered breathing (SDB) is highly prevalent among patients with AF and heart failure and is associated with adverse clinical outcomes. Both the ACC/AHA (5) and ESC guidelines (6) endorse screening for obstructive sleep apnea in patients with AF, particularly when rhythm control strategies are considered. In heart failure (HF) populations, over one-third of patients exhibit SDB with even higher prevalence observed in acute HF (7). Collectively, these guidelines underscore the clinical importance of identifying and characterizing SDB in patients with AF and HF.

Despite the strong theoretical and observational links between OSA and cardiovascular outcomes, real-time, longitudinal monitoring of OSA severity and its dynamic relationship with arrhythmia burden has remained limited in routine clinical practice. Polysomnography, the gold standard for diagnosing OSA, is costly, labour-intensive, and offers only a snapshot assessment. Furthermore, clinical recognition of OSA is often delayed or missed entirely in cardiovascular populations (3,4).

Some implantable cardioverter-defibrillators (ICDs) and cardiac resynchronization therapy defibrillators (CRT-Ds) integrate multiple physiologic sensors to monitor patients remotely for signs of heart failure decompensation, such as the Boston Scientific HeartLogic algorithm. In addition to heart failure metrics, thoracic impedance sensor data has been utilised to estimate an apnea-hypopnea index (AHI), through the AP Scan algorithm. This provides an opportunity to monitor surrogate markers of OSA continuously and non-invasively.

The AP Scan feature on implantable cardiac devices detect breathing patterns by measuring changes in transthoracic electrical impedance that occur with inhalation and exhalation (8). It filters the signal to focus on respiratory frequencies (approximately 6 to 70 breaths per minute, depending on heart rate), effectively removing interference from physical activity and cardiac contractions. From this filtered signal, a respiratory waveform is generated to identify individual breaths and estimate relative tidal volumes. Apnea or hypopnea events are flagged when tidal volume reduces or when no breaths are detected for at least 10 seconds. The total number of such events per hour is reported, correlating well with the apnea-hypopnea index used in sleep studies.

Given the potential clinical impact of OSA on arrhythmia risk, and the availability of novel device-based monitoring technologies, there is a need to better characterize how device-detected sleep apnea correlates with AF burden in a real-world setting. This study aims to evaluate the association between device-derived AHI (via AP Scan) and atrial arrhythmia duration in patients with HeartLogic-enabled CIEDs.

## Methods

### Study Design and Population

We performed a retrospective cohort analysis involving adult patients (aged 18 years and older) who underwent remote monitoring with Boston Scientific HeartLogic-enabled ICD or CRT-D devices at a tertiary hospital’s cardiac implantable electronic device (CIED) clinic between January 1, 2018, and December 31, 2023. Individuals were excluded if they lacked active CIED monitoring or had incomplete follow-up data.

Ethics approval was obtained from the South Western Sydney Local Health District Human Research Ethics Committee (Reference: 2021/STE04860).

In this study, atrial arrhythmia burden was defined as the cumulative daily duration of atrial high-rate episodes (AHRE) detected via implantable cardiac device. AHREs represent periods during which the atrial rate exceeds a device-specific threshold (heart rate ≥175 beats per minute), and to ensure clinical relevance and reduce noise from brief or artifact-driven episodes, we established a minimum threshold of 30 seconds to qualify as an AHRE. This threshold aligns with commonly accepted definitions in device-based arrhythmia monitoring, where shorter episodes may lack diagnostic or prognostic value (9). By summing all qualifying AHREs per 24-hour period, we calculated a mean daily arrhythmia burden (seconds/day), allowing for longitudinal assessment of the temporal relationship between arrhythmia duration and AHI.

### Data Collection

Patient demographics, comorbidities, and medication information were obtained from the electronic medical record and the institutional cardiology database. Device-derived remote monitoring data, including daily HeartLogic and AP Scan readings, were exported from the LATITUDE™ platform. Hospital admissions, along with primary diagnoses and length of stay, were identified through retrospective review of clinical records. All data were curated and managed using the REDCap platform (University of New South Wales).

### Statistical Analysis

A linear mixed-effects model was used to quantify the association between AHI and daily arrhythmia duration (detected atrial high-rate episodes) while accounting for within-patient repeated measures. To further explore the association between OSA severity and arrhythmia duration, we categorized AHI into four groups: none (<5 events/hour), mild (5–15), moderate (15–30), and severe (>30). These thresholds are adapted from standard sleep medicine classifications used in polysomnographic diagnosis and are widely accepted in both clinical and research settings (10). Categorizing AHI values this way allows for comparison of arrhythmia burden across increasing levels of sleep apnea severity, reflecting the progressive physiological stress imposed by more frequent respiratory disturbances. We applied a fixed-effects linear regression to estimate group-wise differences. A multivariable linear mixed-effects model was used to assess the association between patient comorbidities and daily arrhythmia duration, with fixed effects for obesity, chronic kidney disease, stroke, coronary artery disease, diabetes, hyperlipidaemia, hypertension, peripheral vascular disease, chronic obstructive pulmonary disease, obstructive sleep apnea. Patients individual identification was included as a random effect to account for repeated measures. Patients without complete remote monitoring data or inactive HeartLogic indices were excluded.

All statistical analyses were performed using R (version 4.3.2).

## Results

### Patient Characteristics

A total of 400 patients (table 1) were included (mean age 68.6 +/-11.3 years; 76.8% male). The majority of patients had heart failure with reduced ejection fraction (HFrEF, 85.2%), a quarter (24.8%) had a documented diagnosis of obstructive sleep apnea, and one-third (34.3%) were classified as obese (body mass index ≥30), a recognised risk factor for OSA. ICDs were implanted for primary prevention in 59.7% and for secondary prevention in 40.3% of cases. Ischemic cardiomyopathy was the underlying aetiology in 52%, and 48% had non-ischemic cardiomyopathy.

**Table 1:** Patient Characteristics. OSA: Obstructive Sleep Apnea; CKD: Chronic Kidney Disease; AF: Atrial Fibrillation; ACS: Acute Coronary Syndrome; ICD: Implantable Cardioverter Defibrillator

| Patient Characteristic | n (%) |
| --- | --- |
| **Total Patients** | 400 |
| **Mean Age (years)** | 68.6 |
| **Male** | 76.8 |
| **Diabetes Mellitus** | 157 (39.3) |
| **OSA (BMI ≥30)** | 99 (24.8) |
| **Obesity** | 137 (34.3) |
| **CKD** | 92 (23) |
| **AF** | 178 (44.5) |
| **Prior ACS** | 229 (57.3) |
| **Stroke** | 43 (10.8) |
| **Smoking** |  |
| Current Smoker | 57 (14.3) |
| Ex-Smoker | 138 (34.5) |
| Non-Smoker | 178 (44.5) |
| **Cardiomyopathy** | n = 369 |
| Ischaemic Cardiomyopathy | 192 (52) |
| Non-Ischaemic Cardiomyopathy | 177 (48) |
| **ICD indication** | n = 387 |
| Primary Prevention | 231 (59.7) |
| Secondary Prevention | 156 (40.3) |
| **Left Ventricular Function** | n = 366 |
| Normal | 54 (14.8) |
| Mild Impairment | 54 (14.8) |
| Moderate Impairment | 90 (24.5) |
| Severe Impairment | 168 (45.9) |
| **Table 1: Patient Characteristics** |  |
| OSA: Obstructive Sleep Apnea; CKD: Chronic Kidney Disease; AF: Atrial Fibrillation; ACS: Acute Coronary Syndrome; ICD: Implantable Cardioverter Defibrillator |  |

### AHI vs Atrial Arrhythmia Duration

Among the cohort of 126 patients who experienced at least one device-detected atrial high-rate episode (AHRE) lasting ≥30 seconds during the remote monitoring period, we investigated the relationship between sleep-disordered breathing and atrial arrhythmia burden. A linear mixed-effects model, accounting for within-patient variability, demonstrated that each 1-unit increase in daily AHI was associated with a mean increase of 10.55 seconds/day in AHRE duration (95% CI: 3.14 to 17.96, p = 0.005). When using the 30-day averaged AHI to assess longer-term exposure, a 1-unit increase was associated with a 31 second/day increase in arrhythmia duration (95% CI: 19.79 to 41.44, p < 0.001), demonstrating an association between AHI and arrythmia burden.

To explore potential non-linear effects, AHI was also analysed categorically, stratifying patients into none (<5 events/hour), mild (5–15), moderate (15–30), and severe (>30) groups. Compared to those with no significant sleep apnea, patients in all three AHI severity categories had significantly higher daily AHRE durations, with mean increases of approximately 2.8 hours/day for both mild and moderate groups, and 3.0 hours/day for the severe group (all p < 0.001). The apparent discrepancy between the modest per-unit AHI effect and the larger categorical differences likely reflects both the scaling of AHI (where a single-unit increase is clinically minor) and a threshold phenomenon, whereby even mild OSA is associated with a substantial elevation in arrhythmia burden.

In the multivariable mixed-effects model (Figure 1), obstructive sleep apnea was the only comorbidity that remained significantly associated with increased device-detected daily arrhythmia duration after adjustment for other covariates (p = 0.021). None of the other comorbidities evaluated including diabetes, coronary artery disease, stroke, hypertension, obesity, hyperlipidaemia, chronic obstructive pulmonary disease, peripheral vascular disease, and chronic kidney disease demonstrated statistically significant associations, with all confidence intervals crossing the null. These findings suggest that the association between OSA and arrhythmia burden is independent of other common cardiovascular and systemic conditions.

**Figure 1:**
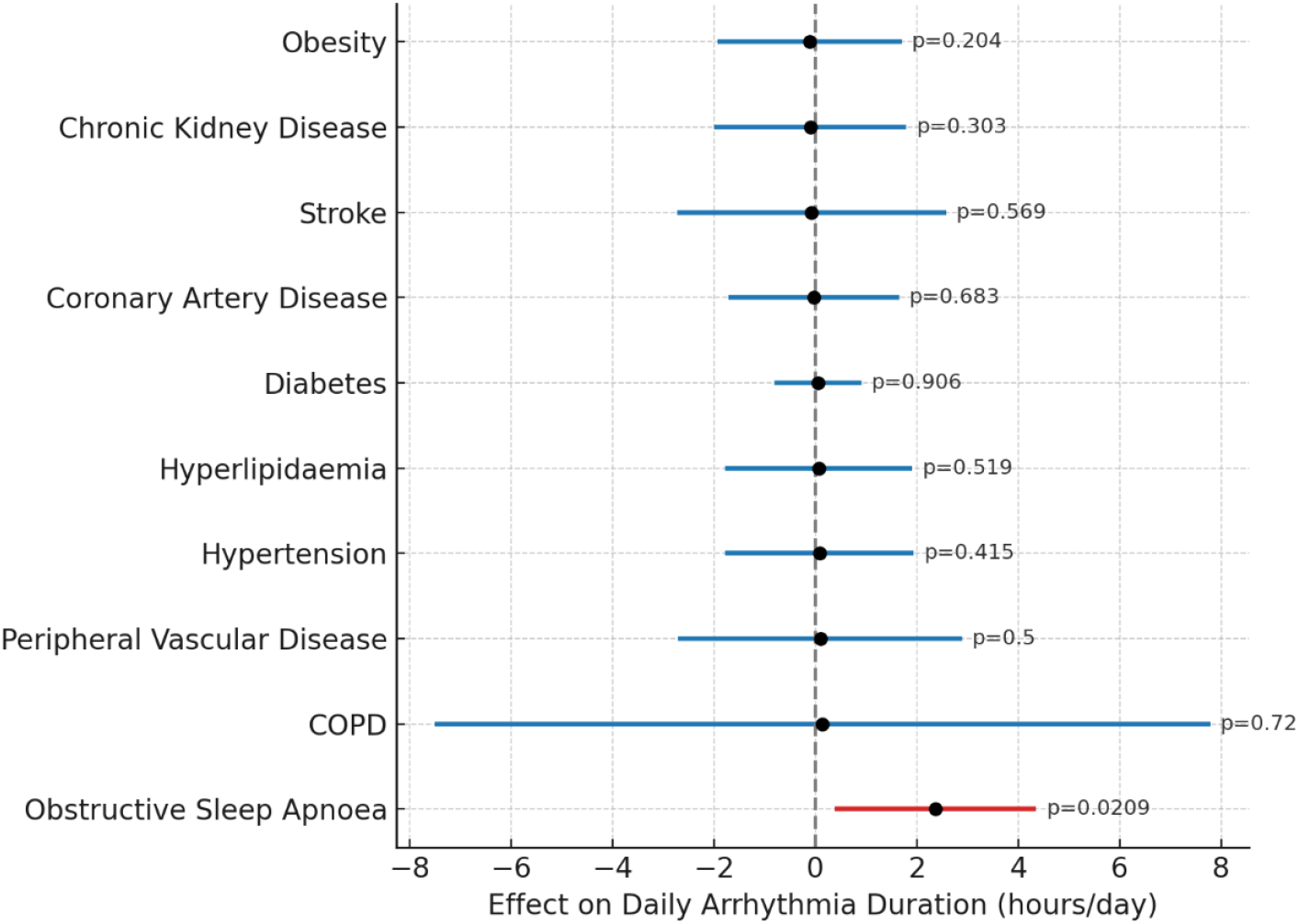
Patient Comorbidities and Device-Detected Arrhythmia Burden.

## Discussion

In this study, we explored the association between device-detected AHI and atrial arrhythmia burden in a cohort of patients with CIEDs. Utilising daily device-captured data, we observed a significant, dose-dependent associated between AP Scan-derived AHI and arrhythmia duration. Each 1-unit increase in daily AHI was associated with an average increase of 10.55 seconds/day in atrial high-rate episode (AHRE) duration (p = 0.05). The association was even stronger when using a 30-day averaged AHI, where each unit increase corresponded to an additional 31-second/day rise in arrhythmia burden (p < 0.001). For example, a 10-unit increase in AHI corresponded to an estimated increase of 310 seconds/day in AHRE duration. Stratified analyses revealed increased arrhythmia duration across mild, moderate, and severe AHI categories, with increases of 2.8 to 3.0 hours/day compared to patients with no OSA.

The differences observed between the continuous and categorical analyses warrant clarification. The per-unit estimates (10–30 seconds/day per AHI unit) appear modest when considered in isolation, however, over the span of typical clinical AHI ranges (eg. differences of 10–20 units between patients), these increments may accumulate to several minutes or hours of additional arrhythmia burden per day. By contrast, the categorical analysis suggests a threshold effect, with the presence of OSA whether mild, moderate, or severe being associated with a marked increase in arrhythmia duration, but little incremental increase across higher severity of apnea. This pattern is consistent with potential non-linear pathophysiological effects of sleep-disordered breathing on atrial substrate and raises the hypothesis that even mild OSA may have clinically significant impacts in the heart failure population.

Atrial arrhythmias, particularly device detected atrial high-rate, are increasingly recognized as clinically meaningful markers of cardiovascular risk. They may reflect underlying atrial pathology and are associated with stroke, heart failure progression, and all-cause mortality (9). Device-detected AHREs provide an opportunity for risk stratification and potential early intervention, particularly when measured longitudinally in high-risk populations.

In a multivariable mixed-effects model, other comorbidities such as diabetes, coronary artery disease, stroke, hypertension, obesity, hyperlipidaemia, chronic obstructive pulmonary disease, peripheral vascular disease, and chronic kidney disease did not demonstrate a significant association with increased device-detected arrhythmia burden.

Atrial fibrillation is the most prevalent sustained arrhythmia globally, characterized by uncoordinated atrial activity resulting in loss of atrioventricular synchrony and impaired cardiac efficiency. Its pathogenesis involves a complex interplay of electrical and structural remodelling, including atrial fibrosis, chamber dilation, and infiltration of inflammatory cells. These changes are further influenced by autonomic nervous system imbalance and oxidative stress, all of which create an atrial substrate vulnerable to arrhythmia initiation and maintenance (11,12). Over time, AF becomes self-perpetuating where repetitive episodes further promote electrical and structural changes.

Obstructive sleep apnea is common and is increasingly recognized as a potent arrhythmogenic factor. OSA is characterized by repetitive upper airway obstruction during sleep, resulting in intermittent hypoxia, hypercapnia, large negative intrathoracic pressure swings, and recurrent arousals. These disturbances activate a cascade of physiological responses that adversely affect the cardiovascular system. Intermittent hypoxia triggers the generation of reactive oxygen species and upregulation of hypoxia-inducible pathways, promoting inflammation and myocardial fibrosis (13). Simultaneously, negative intrathoracic pressures during obstructed breathing episodes increase atrial wall stretch and chamber distension, exacerbating mechanical stress on atrial tissue (14). These changes are compounded by sympathetic nervous system activation and transient surges in blood pressure and heart rate, which contribute to both substrate and trigger mechanisms underlying atrial fibrillation (15,16).

The mechanistic link between OSA and AF has been well documented in both experimental and clinical studies. Animal models have demonstrated that intermittent hypoxia can lead to atrial fibrosis, autonomic imbalance, and altered calcium handling, changes that closely mirror human AF pathophysiology (3). In the Sleep Heart Health Study, OSA severity was directly associated with increased AF prevalence (17). Mehra et al. demonstrated that patients with moderate to severe OSA had a four-fold higher risk of AF compared to those without sleep-disordered breathing (18).

Furthermore, in a detailed electrophysiological and electroanatomic study, Dimitri et al. investigated atrial remodeling in patients with moderate-to-severe OSA undergoing catheter ablation for paroxysmal AF. Compared to matched controls without OSA, affected individuals demonstrated significant atrial structural and electrical abnormalities, including bi-atrial enlargement, reduced atrial voltage, areas of electrical silence, conduction slowing, and prolonged sinus node recovery times. Furthermore, patients with OSA exhibited a higher burden of complex electrograms, suggesting widespread conduction heterogeneity. These findings were observed in the absence of differences in atrial effective refractory periods, implicating OSA-induced atrial remodeling as a potential substrate for both the development and maintenance of AF (19).

Despite these insights, real-world monitoring of OSA in patients with cardiovascular disease has been constrained by the limitations of traditional polysomnography (PSG), which is costly, labor-intensive, and only captures a snapshot in time. Implantable cardiac electronic devices (CIEDs), equipped with AP Scan functionality, represent a paradigm shift by enabling continuous, passive estimation of AHI using thoracic impedance. This allows longitudinal assessment of nightly respiratory disturbance patterns without patient input or disruption to routine care (20,21).

The study findings are consistent with and build upon prior studies examining the relationship between device-detected sleep apnea and atrial arrhythmias. In the DASAP-HF study, Boriani et al. reported that patients with a respiratory disturbance index (RDI) ≥32/h detected via ICDs had a significantly increased risk of AF (22). The DEDICATES study further demonstrated that higher AHI values, detected through remote monitoring of CRT and ICD devices, correlated with increased atrial tachyarrhythmia burden (23,24). Similarly, Mazza et al. found that severe sleep apnea detected via pacemaker telemetry (RDI ≥30) predicted a 2.8-fold increased risk of new-onset AF (25). While most of these studies used binary outcomes such as AF presence or recurrence, our study expands upon these results by quantifying arrhythmia burden on a continuous scale and demonstrating a linear relationship with rising AHI.

Moreover, previous validation studies have confirmed the utility of device-based respiratory monitoring for long-term risk stratification, despite some limitations in accuracy when compared with gold-standard PSG. Barbieri et al. reported a moderate correlation between AP Scan-derived AHI and PSG values (r = 0.41), suggesting that while device-based metrics may overestimate absolute AHI values, they remain valuable for tracking temporal trends and identifying high-risk patients (8). A meta-analysis evaluated the use of implantable cardiac devices (Boston Scientific, ELA Medical, Sorin/LivaNova, Medtronic) for sleep apnea detection using impedance-derived respiratory indices. Across 16 studies, device-based monitoring showed good diagnostic performance for severe OSA (AHI ≥30), with pooled sensitivity and specificity of 78% and 79%, respectively (26). These findings support the use of cardiac devices for opportunistic OSA screening in patients with atrial fibrillation.

Importantly, this study is hypothesis-generating. First, it highlights a potential association between even mild OSA and increased arrythmia burden, raising the possibility that subclinical OSA contributes to arrhythmogenic substrate. Second, it demonstrates the feasibility of continuous device-based OSA monitoring as a clinical tool for dynamic risk stratification in patients with existing CIEDs. In conclusion, this study suggests a potential association between OSA, as detected by AP Scan-derived AHI from implantable cardiac devices, and atrial arrhythmia burden. This association was consistent across daily and averaged AHI values and persisted after accounting for intra-individual variability. These findings should be interpreted as exploratory and hypothesis-generating rather than causal. Future prospective trials are warranted to clarify whether device-detected AHI directly contributes to arrythmia burden and whether targeted interventions based on device-detected AHI can reduce AF burden and improve clinical outcomes.

### Limitations

This study has several limitations that warrant consideration. First, as a retrospective analysis, it is inherently subject to residual confounding and does not permit definitive causal inference between device-detected increases in AHI and clinical outcomes such as atrial arrythmia burden. The patient population, predominantly older men with HFrEF, may not represent the broader AF/OSA population, thereby limiting external validity. The single-centre design may limit the generalisability of these findings to broader or more diverse populations. The use of AP Scan-derived AHI represents a surrogate measure of sleep apnea burden which is not validated as a diagnostic test and may not fully capture the complexity or severity of sleep-disordered breathing compared to formal polysomnography, however AP Scan-derived AHI may be used as a screening test for patients with implanted compatible devices and be useful in identifying high risk patients with undiagnosed sleep apnea in an already high risk group. Additionally, device-detected atrial high-rate episodes (AHREs) may include atrial flutter or other atrial tachyarrhythmias, and are not specific to atrial fibrillation. As electrograms (EGMs) for individual AHRE events were not available for review, we were unable to definitively classify the arrhythmia subtype for each episode. This lack of rhythm validation introduces potential misclassification bias in the interpretation of arrhythmia burden. Furthermore, data on continuous positive airway pressure (CPAP) therapy and formal sleep study–based OSA diagnoses were not available and therefore not included in the analysis. These findings should be interpreted as hypothesis-generating, and future prospective, multicentre studies incorporating comprehensive sleep diagnostics are warranted to validate and expand upon these observations.

## Conclusion

Our study demonstrates an association between device detected OSA and increased atrial arrhythmia burden in heart failure patients monitored with multiparametric devices. These findings support prior observations suggesting a relationship between obstructive sleep apnea and atrial fibrillation and offer novel insights through the use of continuous longitudinal data from implantable device telemetry. While the observational nature of this study precludes causal inference, the results highlight a potential role for incorporating sleep-disordered breathing surveillance into remote cardiac monitoring. This approach may help identify patients at increased risk of arrhythmia or heart failure progression and supports further investigation into whether tailored interventions based on device-detected OSA could improve outcomes.

## Data Availability

All data produced in the present study are available upon reasonable request to the authors

## References

1. Yeghiazarians Y, Jneid H, Tietjens JR, Redline S, Brown DL, El-Sherif N, et al. Obstructive Sleep Apnea and Cardiovascular Disease: A Scientific Statement From the American Heart Association. Circulation. 2021 July 20;144(3).

2. Basner RC. Cardiovascular Morbidity and Obstructive Sleep Apnea. N Engl J Med. 2014 June 12;370(24):2339–41.

3. Zhang L, Hou Y, Po SS. Obstructive Sleep Apnoea and Atrial Fibrillation. Arrhythmia Electrophysiol Rev. 2015;4(1):14.

4. Mehra R, Chung MK, Olshansky B, Dobrev D, Jackson CL, Kundel V, et al. Sleep-Disordered Breathing and Cardiac Arrhythmias in Adults: Mechanistic Insights and Clinical Implications: A Scientific Statement From the American Heart Association. Circulation. 2022 Aug 30;146(9).

5. Joglar JA, Chung MK, Armbruster AL, Benjamin EJ, Chyou JY, Cronin EM, et al. 2023 ACC/AHA/ACCP/HRS Guideline for the Diagnosis and Management of Atrial Fibrillation: A Report of the American College of Cardiology/American Heart Association Joint Committee on Clinical Practice Guidelines. Circulation. 2024 Jan 2;149(1).

6. Van Gelder IC, Rienstra M, Bunting KV, Casado-Arroyo R, Caso V, Crijns HJGM, et al. 2024 ESC Guidelines for the management of atrial fibrillation developed in collaboration with the European Association for Cardio-Thoracic Surgery (EACTS). Eur Heart J. 2024 Sept 29;45(36):3314–414.

7. McDonagh TA, Metra M, Adamo M, Gardner RS, Baumbach A, Böhm M, et al. 2021 ESC Guidelines for the diagnosis and treatment of acute and chronic heart failure. Eur Heart J. 2021 Sept 21;42(36):3599–726.

8. Barbieri F, Dichtl W, Heidbreder A, Brandauer E, Stefani A, Adukauskaite A, et al. Sleep apnea detection by a cardiac resynchronization device integrated thoracic impedance sensor: A validation study against the gold standard polysomnography. Fukumoto Y, editor. PLOS ONE. 2018 Apr 6;13(4):e0195573.

9. Sagris D, Georgiopoulos G, Pateras K, Perlepe K, Korompoki E, Milionis H, et al. Atrial High‐Rate Episode Duration Thresholds and Thromboembolic Risk: A Systematic Review and Meta‐Analysis. J Am Heart Assoc. 2021 Nov 16;10(22):e022487.

10. Douglas JA, Chai-Coetzer CL, McEvoy D, Naughton MT, Neill AM, Rochford P, et al. Guidelines for sleep studies in adults – a position statement of the Australasian Sleep Association. Sleep Med. 2017 Aug;36:S2–22.

11. Nattel S, Harada M. Atrial Remodeling and Atrial Fibrillation. J Am Coll Cardiol. 2014 June;63(22):2335–45.

12. Andrade J, Khairy P, Dobrev D, Nattel S. The Clinical Profile and Pathophysiology of Atrial Fibrillation: Relationships Among Clinical Features, Epidemiology, and Mechanisms. Circ Res. 2014 Apr 25;114(9):1453–68.

13. Lavie L. Oxidative stress in obstructive sleep apnea and intermittent hypoxia – Revisited – The bad ugly and good: Implications to the heart and brain. Sleep Med Rev. 2015 Apr;20:27–45.

14. Bradley TD, Floras JS. Obstructive sleep apnoea and its cardiovascular consequences. The Lancet. 2009 Jan;373(9657):82–93.

15. Somers VK, Dyken ME, Mark AL, Abboud FM. Sympathetic-Nerve Activity during Sleep in Normal Subjects. N Engl J Med. 1993 Feb 4;328(5):303–7.

16. Iwasaki Y ki, Nishida K, Kato T, Nattel S. Atrial Fibrillation Pathophysiology: Implications for Management. Circulation. 2011 Nov 15;124(20):2264–74.

17. Gami AS, Pressman G, Caples SM, Kanagala R, Gard JJ, Davison DE, et al. Association of Atrial Fibrillation and Obstructive Sleep Apnea. Circulation. 2004 July 27;110(4):364–7.

18. Mehra R, Benjamin EJ, Shahar E, Gottlieb DJ, Nawabit R, Kirchner HL, et al. Association of Nocturnal Arrhythmias with Sleep-disordered Breathing: The Sleep Heart Health Study. Am J Respir Crit Care Med. 2006 Apr 15;173(8):910–6.

19. Dimitri H, Ng M, Brooks AG, Kuklik P, Stiles MK, Lau DH, et al. Atrial remodeling in obstructive sleep apnea: Implications for atrial fibrillation. Heart Rhythm. 2012 Mar;9(3):321–7.

20. Boehmer JP, Hariharan R, Devecchi FG, Smith AL, Molon G, Capucci A, et al. A Multisensor Algorithm Predicts Heart Failure Events in Patients With Implanted Devices. JACC Heart Fail. 2017 Mar;5(3):216–25.

21. Ziegler PD, Koehler JL, Mehra R. Comparison of continuous versus intermittent monitoring of atrial arrhythmias. Heart Rhythm. 2006 Dec;3(12):1445–52.

22. Boriani G, Diemberger I, Pisanò ECL, Pieragnoli P, Locatelli A, Capucci A, et al. Association between implantable defibrillator‐detected sleep apnea and atrial fibrillation: The DASAP‐HF study. J Cardiovasc Electrophysiol. 2022 July;33(7):1472–9.

23. Gwag HB, Park Y, Lee SS, Kim JS, Park KM, On YK, et al. Rationale, design, and endpoints of the ‘DEvice-Detected CArdiac Tachyarrhythmic Events and Sleep-disordered Breathing (DEDiCATES)’ study: Prospective multicenter observational study of device-detected tachyarrhythmia and sleep-disordered breathing. Int J Cardiol. 2019 Apr;280:69–73.

24. Gwag HB, Park Y, Lee SS, Kim JS, Park KM, On YK, et al. Rationale, design, and endpoints of the ‘DEvice-Detected CArdiac Tachyarrhythmic Events and Sleep-disordered Breathing (DEDiCATES)’ study: Prospective multicenter observational study of device-detected tachyarrhythmia and sleep-disordered breathing. Int J Cardiol. 2019 Apr;280:69–73.

25. Mazza A, Bendini MG, De Cristofaro R, Lovecchio M, Valsecchi S, Boriani G. Pacemaker-detected severe sleep apnea predicts new-onset atrial fibrillation. EP Eur. 2017 Dec 1;19(12):1937–43.

26. Ben Messaoud R, Khouri C, Pépin JL, Cracowski JL, Tamisier R, Barbieri F, et al. Implantable cardiac devices in sleep apnoea diagnosis: A systematic review and meta-analysis. Int J Cardiol. 2022 Feb;348:76–82.

